# A Wearable Multi-modal Sensor Array for Continuous Cuffless Blood Pressure Estimation

**DOI:** 10.64898/2026.01.25.26344788

**Authors:** John Rattray, Bright Nnadi, Sampath Rapuri, Carl Harris, Francesco Tenore, Charlene Gamaldo, Robert D Stevens, Ralph Etienne-Cummings

**Affiliations:** Whiting School of Engineering, Johns Hopkins University, Baltimore, USA; Research and Exploratory Development Dept, Johns Hopkins Applied Physics Laboratory, Laurel, USA; Johns Hopkins School of Medicine, Johns Hopkins University, Baltimore, USA; School of Medicine, Johns Hopkins University, Baltimore, USA

**Keywords:** wearable, continuous, blood pressure, non-invasive, multi-modal, multi-site, Pulse Arrival Time, Pulse Transit Time, Pre-ejection Period

## Abstract

Blood pressure (BP) measurement is crucial for medical care, yet existing BP methods are either invasive, tethered, or suffer from low temporal resolution. Non-invasive continuous BP estimation thus remains a significant challenge. To address these challenges, this work presents a novel, non-invasive, multi-modal sensor designed for continuous blood pressure estimation using multiple biosignal modalities as feature inputs. From these input data, we extract cardiovascular timing intervals (e.g., pulse arrival time), which serve as key features for BP regression models, enabling continuous, non-invasive BP monitoring. We validate our algorithm with 16 healthy subjects using standard blood pressure cuff readings as ground truth. Our wearable, non-invasive multimodal and multinodal sensor array for integrated computation (MOSAIC) demonstrated promising performance and was able to predict systolic and diastolic BP across all study subjects with a MAE of 5.31 ± 7.32 mmHg and 4.27 ± 2.35 mmHg, respectively.

## I. Introduction

Hypertension is one of the most pervasive and costly chronic conditions worldwide, affecting almost 1 in 2 adults aged 18 and over and ranging from about 1 in 5 young adults (18-39) to nearly 3 in 4 adults aged 60 and over in the United States alone according to the CDC [https://www.cdc.gov/nchs/products/databriefs/db364.htm].

Despite its prevalence and the well-established complications associated with poor management of the disease, such as stroke, cardiovascular disease, and organ damage, less than a quarter of hypertensive individuals maintain adequate control over their blood pressure. About 70% of the hypertensive population requires a combination of at least two antihypertensive medications to reach target levels [13]. This alarming gap in management imposes substantial economic burden, with direct medical costs and productivity losses exceeding $131 billion annually in the United States [1]. The inadequacy of current monitoring technologies represents a critical barrier to effective hypertension management, as these systems fail to provide accurate, continuous, and user-friendly assessments of blood pressure in real-world settings.

Blood pressure is a dynamic biosignal regulated in part by the autonomic nervous system (ANS), and its continuous monitoring offers critical insights into cardiovascular health, disease progression, and effects of therapy. The gold standard for continuous blood pressure measurement is arterial catheterization, in which a cannula (arterial line or A-line) is inserted directly into the arterial lumen to enable beat-to-beat pressure monitoring with high temporal resolution. While this approach is unparalleled in its accuracy and enables observation of blood pressure variability throughout each cardiac cycle, it introduces significant clinical risks including bleeding, arterial damage, infection, and hematoma formation. Consequently, Aline monitoring is restricted to intensive care units and operative settings where the clinical benefits justify the inherent risks and resource requirements. A noninvasive alternative, the sphygmomanometer, represents the most widely adopted approach for blood pressure measurement both in clinical and in-home settings. These devices offer greater accessibility and eliminate procedural risks, but suffer from fundamental limitations that compromise their clinical utility. Moreover, an improved blood pressure monitoring system, particularly when accessible outside a brick-and-mortar facilities, has the potential to enable faster clinical response, support timely treatment adjustment, and expand access to effective hypertension management even outside a hospital setting. Oscillometric measurements exhibit systematic errors of *±*6 mmHg for both systolic and diastolic readings [2]. Moreover, obtaining accurate readings requires strict adherence to measurement protocols: patients must remain unstimulated, seated, and at rest for 7 minutes or longer [3] with proper cuff positioning and arm support. These requirements limit the temporal resolution, making it impossible to capture blood pressure variability throughout daily activities or detect acute hypertensive episodes. In contrast, to emerging approaches such as MOSAIC that offer significant advantages, including continuous blood-pressure estimation, reduced wearability discomfort, improved sensitivity to high blood pressure variations, enhanced data accessibility; features that addresses limitations with conventional oscillometric devices and provides meaningful improvements to real-world hypertension monitoring.

Recent advances in wearable health technologies have opened new avenues for noninvasive, continuous monitoring of physiological signals. These devices promise to revolutionize chronic disease management by providing longitudinal health data without disrupting normal behavior patterns. However, most commercially available devices rely on single-site measurements typically taken at the wrist using tethered photoplethysmography (PPG) and accelerometer sensors which are particularly susceptible to motion artifacts, poor peripheral perfusion, and limited anatomical relevance for central blood pressure estimation. As such, current wearable blood pressure monitoring solutions face substantial technical challenges that limit their clinical validity and practicality. Furthermore, existing algorithms often lack robustness across diverse patient populations, with reduced accuracy in individuals with diabetes and peripheral vascular disease. These limitations result in unreliable blood pressure estimation that fail to meet regulatory standards in clinical settings. By combining signals from chest and peripheral (finger) sites, this work’s approach captures both cardiac electrical activity and arterial pressure wave propagation, providing rich temporal and spatial information about hemodynamic function. The overarching goal of this research is to bridge the gap between invasive precision and non-invasive practicality for blood pressure measurement in both clinical and everyday environments.

This research study introduces the Multimodal and multinodal Sensor Array for Integrated Computation (MOSAIC) system, a novel non-invasive wearable system designed to address the fundamental limitations of existing blood pressure monitoring technologies. MOSAIC employs a custom-designed distributed sensor architecture that simultaneously captures ECG, PPG, and accelerometer signals from strategic anatomical locations, Bluetooth Low Energy (BLE) data transmission, and mobile computation to enable comprehensive characterization of cardiovascular dynamics. From detailed data collection protocols, advanced signal processing techniques, and machine learning model development, this work seeks to establish the feasibility and clinical validity of multisite, multi-modal sensing for blood pressure management, ultimately contributing to more effective interventions for one of the most critical public health challenges of our time.

## II. Methods

The development of the MOSAIC system emerged from limitations identified during prior research monitoring output of severe brain injury (SBI) patients [14]. The MOSAIC system adopts elements of conventional wearable design including form factor, rechargability, device interchangeability, low-latency and low power strategies while challenging others such as single-site measurements. This custom-engineered solution comprises an array of reconfigurable, synchronized sensor nodes that continuously acquire multi-modal biosignals from distributed body locations and which wirelessly transmit data to a central device for real time visualization and storage. This system integrates four complementary modalities selected based on their physiological relevance to autonomic function and optimal power-size characteristics: **mechanical sensing** via Micro-Electro-Mechanical-System (MEMS) accelerometry for motion and vibration signal; **biopotential measurement** for cardiac electrophysiology (ECG) capturing electrical activity from excitable tissues (resting potential ~ −70 mV, action potential peaks *~* +40 mV), through surface recordings typically measure of 1 mV or less requiring amplification; **electrodermal activity** (EDA) monitoring of sympathetic nervous system activation through skin conductance changes; **photoplethysmography** (PPG) using optical volumetric phlethysmograhpy to capture cardiovascular pulsatile dynamics.

### A. MOSAIC Hardware Design

The MOSAIC sensor employs a novel three-layer vertically stacked PCB configuration (40mm x 13mm x 0.4mm) designed for unobtrusive daily wear (Figure 2). The top layer houses the nRF52832 microcontroller (MCU) (MDBT42Q module), user interface components (dual-color LEDs, pushbuttons) and a 40mAh rechargeable battery with 3.3V voltage regulation. The middle layer is comprised of an AD8232-based ECG analog front end with 100 gain, an EDA circuit measuring skin conductance (2-20*µ*S), and a 4-channel multiplexer that enables shared electrode use between ECG and EDA blocks under MCU control. The bottom layer contains the MAX30101 optical sensor for infrared PPG acquisition and the ADXL343 accelerometer. The system features embedded snap connectors spaced 40mm apart on the underside of the middle PCB for direct attachment of 30mm x 24mm Ag/AgCl wet electrodes, eliminating the need for conventional lead wires.A custom 3D-printed enclosure protects components while maintaining wearable comfort. The compact design necessitated a specialized charging dock with push-on, spring loaded (POGO) pin contacts which simultaneously charges three devices.

**Fig. 1.**
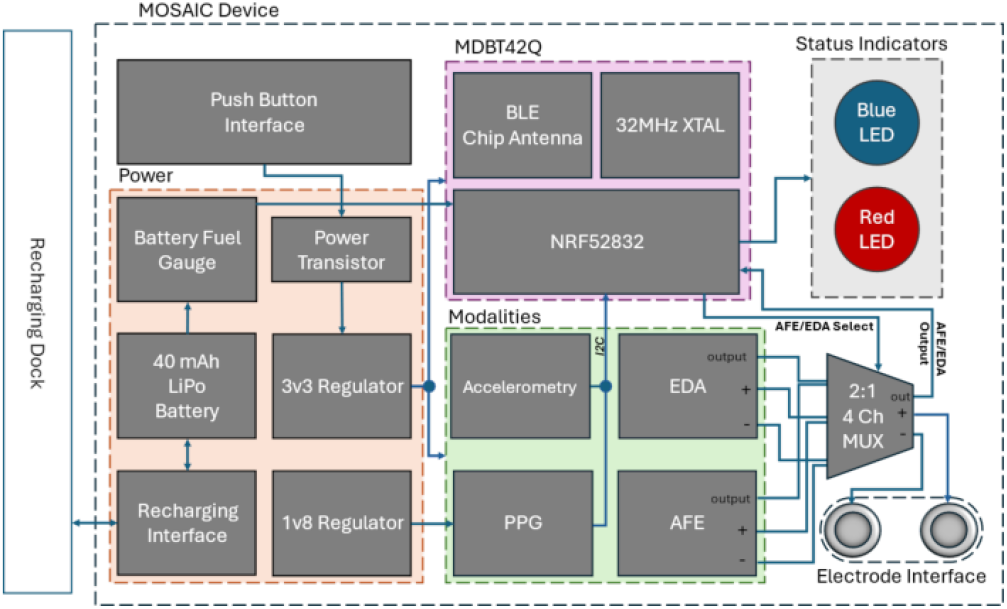
A block diagram showing the main hardware blocks of the MOSAIC device

**Fig. 2.**
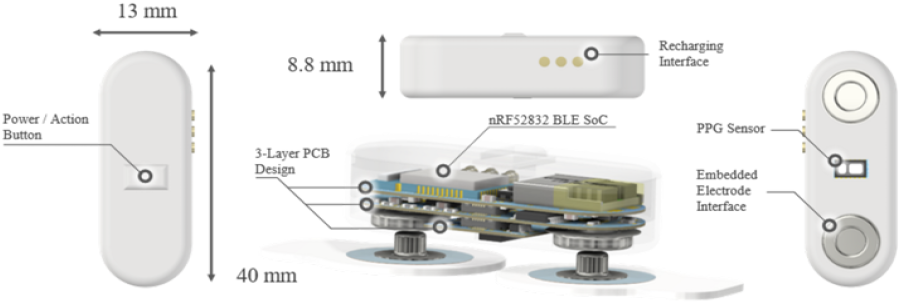
A 3D rendering of the fully assembled MOSAIC device. The internal componentry is visualized through a translucent body showing the 3-layer PCB design, embedded electrodes, and battery

Synchronicity of acquired physiological data from each node is a defining characteristic of the MOSAIC system. Unlike single-site devices, the MOSAIC system performs simultaneous measurements from multiple body locations, reducing measurement errors through cross-referencing and advanced signal characterization across different body regions. Low-power wireless sensor networks present synchronization challenges due to limited resources, variable communication latency, and constrained bandwidth. We employed the Flooding Time Synchronization Protocol (FTSP), which achieves synchronization by timestamping a single radio message at both sender and receiver [4]. Our implementation utilized Nordic Semiconductor’s TimeSync library, modified for our research objective. Accurate timekeeping relied on the nRF’s programmable peripheral interconnect (PPI), enabling autonomous hardware interactions between radios, GPIOs, clocks, and registers independent of CPU intervention. Each node maintains a free-running 16 MHz timer synchronized to a timing master that transmits synchronization packets at configurable intervals. Receiver nodes update their local timers by applying correction offsets based on received timestamps [5]. The synchronization accuracy was validated by connecting the timer’s LSB to a GPIO output, enabling observation of oscillation difference between devices. This achieved a maximum synchronization error of 200ns, over 4 orders of magnitude smaller than the device’s highest sampling frequency (250 Hz).

### B. MOSAIC Firmware and Data collector architecture

The firmware developed for the MOSAIC system was written in C on the Zephyr RTOS and employs a finite state machine (FSM) with seven distinct operational states to manage device behavior systematically, as presented in Table I. Each Node captures multiple physiological signals including electrodermal activity, photoplethysmography, tri-axial accelerometry, and biopotentials from different anatomical sites. This multi-modal, multi-nodal approach provides continuous insight into cardiovascular, respiratory, and autonomic processes. Coherent temporal alignment across modalities is achieved via timestamped data packets and a unified sampling framework, enabling synchronized physiological monitoring with low power consumption and latency. Bluetooth Low Energy (BLE) was used as the wireless protocol for transmitting sensor data to the host system, operating in the 2.4 GHz ISM band.

**TABLE I.** Finite State Machine (FSM) transition table for MOSAIC firmware.

| Current State | Event / Input | Next State |
| --- | --- | --- |
| Off | Button pressed | Start-up |
| Start-up | Peripherals initialized | Advertising |
| Advertising | BLE connection | Connected |
| Connected | Host: provision | Provisioning |
| Provisioning | Config complete | Ready |
| Ready | Start command received | Collecting Data |
| Collecting Data | Stop command | Ready |
| Any | Disconnection event | Off |

Data were transmitted in structured packets optimized for BLE’s limited 244-byte payload capacity detailed in Figure 3. Each packet contained a compact header with a device identifier, datatype label, and timestamp, followed by the corresponding sensor data. Buffers for each sensor modality were managed independently to prevent overflow and to ensure that transmission timing reflected actual sampling intervals. This architecture maximized available bandwidth, supported accurate temporal reconstruction during analysis, and enabled the host application to visualize and store data in real time. The design achieved efficient, synchronized data transfer despite the inherent constraints of low-energy wireless communication.

**Fig. 3.**
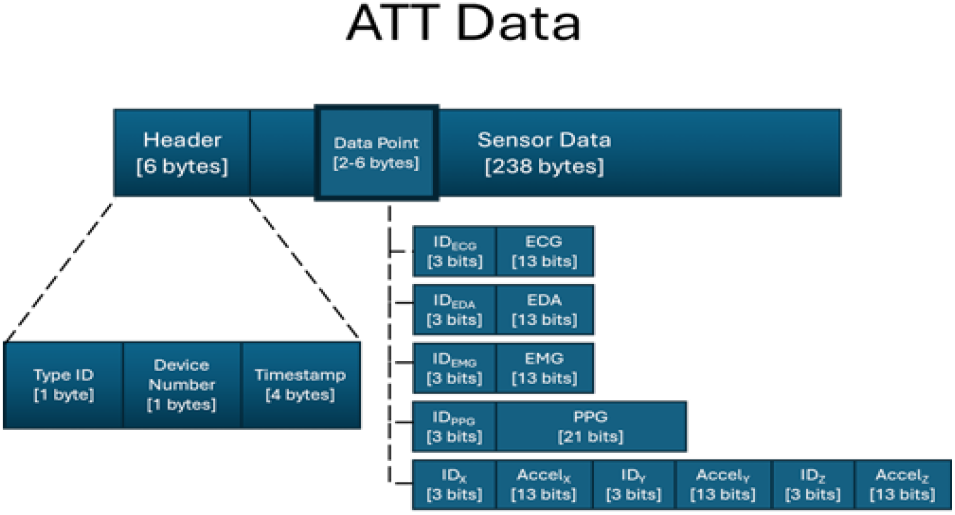
Visualization of the 244 ATT data block and the breakdown of the data packet for encapsulating device metadata, timestamp, and sensor data

**Fig. 4.**
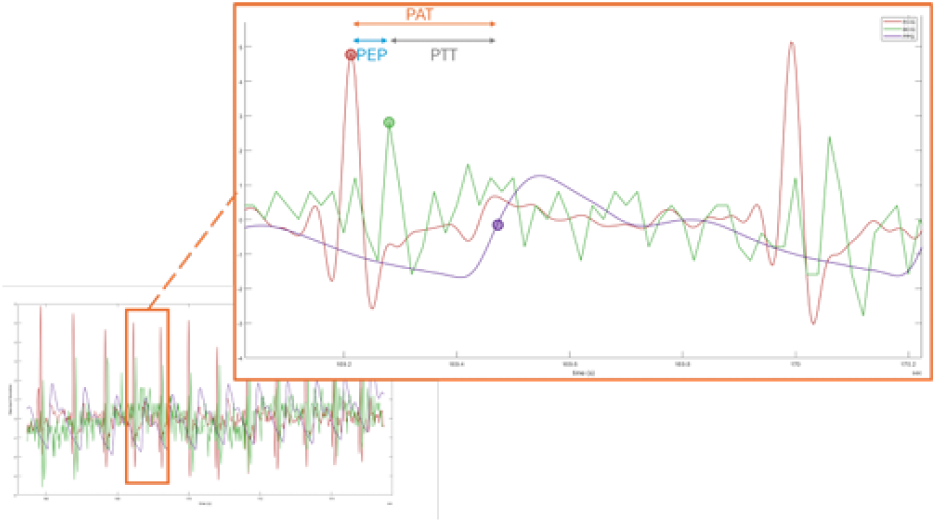
A 10-second window of ECG (red), BCG (green), and PPG (purple) extracted from participant 12. (Magnified) A 1-second selection detailing the fiducial points used to measure PAT, PEP, and PTT

### C. Signal Processing

A MATLAB-based application was developed to streamline all processes of data capture, device connectivity, and real-time visualization for the MOSAIC system.

To improve the biosignal quality, the MOSAIC sensors employ hardware and algorithmic filtering. The first stage of noise reduction is achieved through analog filtering on the device to remove powerline interference and radio frequency artifacts.The next stage is achieved in Software preprocessing that includes domain-specific filtering for each modality. High-frequency noise in ECG, PPG, and BCG signals is attenuated using a 10 Hz low-pass filter. PPG data additionally undergo a 3 Hz low-pass filter to preserve waveform morphology by removing 400 Hz LED-induced artifacts. BCG acceleration signals are high-pass filtered at 5 Hz to remove slow baseline drift.

Pulse arrival time (PAT) is derived as the time interval between the ECG R-peak and a fiducial point on the peripheral PPG waveform, while the pre-ejection period (PEP) is the measure as the time difference between the ECG R-peak and BCG J-peak, which corresponds to the onset of the ventricular blood ejection into the aorta represented in figure (4) [8], [10]. We model the relationship between pulse arrival time (PAT) and blood pressure, a connection extensively reported in prior work [6], [7], [11]. We also derive pulse transit time (PTT) from the following equation:

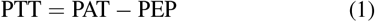

where PTT is the pulse transit time, and PEP is the pre-ejection period. On the filtered ECG data, we select R-peaks using the HRV Analysis Toolkit [9]. The software successfully detected the majority of R peaks within the ECG waveforms but was unable to identify R-peaks in regions of low signal to noise ratio. In these cases, we manually annotate the R peaks. This same process was applied to the detection of upstroke points in the PPG waveform, utilizing the PPG’s first derivative. Lastly, the J-peak of the BCG waveform is extracted by observing the local maxima of the Z-axis of the accelerometer around a local window of 300 ms surrounding each R-peak.

### D. Research Study

A Johns Hopkins Institutional Review Board approved study was conducted with sixteen adult participants with no neurological or psychiatric impairments and the ability to climb four flights of stairs unassisted. The cohort (31% female) had a mean age of 35*±*9 years and a BMI of 27 *±*4; six participants met the criteria for hypertension. Data were collected from three MOSAIC devices during seven measurement sessions: baseline, deep breathing, Valsalva Maneuver, active standing, mental arithmetic, breath hold, and stair climbing. The MOSAIC devices were placed on the chest between the 4th and 5th intercostal space, on the index finger, and between the middle and ring fingers on the right hand. Blood pressure was obtained at 15 distinct timepoints using a standard cuff located on the left arm.

### E. Modeling

For each cuff measurement, 30 seconds of preceding PAT data derived from the MOSAIC devices on the chest and index finger were extracted.The mean PAT of that period was used as the only feature during training of a univariate linear regression model. Model performance was evaluated using leave-one-out cross-validation (LOOCV), whereby the model is trained on all but one sample and evaluated on the withheld sample (this process is repeated until the model is evaluated on each sample and errors are averaged across all evaluations) (consider having a more concise form of description like this). The model was then applied to the remaining data collected to produce continuous BP estimation over the course of the 7-session research study.

## III. Results

Figure 5 reports mean absolute error (MAE) and standard deviation (STD) achieved in applying a linear regression model to fit each participant’s systolic and diastolic BP readings to their corresponding PAT measurements. The models were collectively able to predict systolic and diastolic BP with a MAE of 5.31 ± 7.32 mmHg and 4.27 ± 2.35 mmHg respectively. Three measurement sessions were discarded from participant 16’s data due to low PPG SNR and indiscernible fiducial features needed for PAT extraction. Table II shows that the MOSAIC system provides competitive systolic blood pressure estimation compared to three recent cuffless methods and achieves the most precise diastolic prediction, despite an average MAE 0.31mmHg higher than the group average. Unlike single-device systems in prior research which require overt user actions and interrupt continuous monitoring, the MOSAIC system’s multi-device design enables uninterrupted, continuous blood pressure estimation without special user maneuvers, offering a unique advantage over current state-of-the-art approaches. The results of the continuous systolic BP estimation are demonstrated for a single participant in Figure 7. This participant was pre-hypertensive, had a BMI categorized as healthy and did not have any major health conditions. The largest decrease in PAT and corresponding increase in BP can be seen during the active stand and stair climb tests which is expected as these tests involved the most physical activity. The other sessions with notable increases in BP were the Valsalva maneuver and the breath hold.

**TABLE II.**
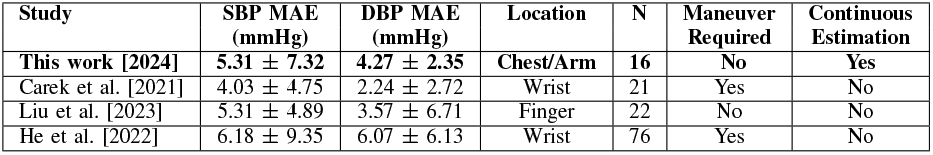
Comparison with state-of-the-art cuffless BP estimation methods.

| Study | SBP MAE (mmHg) | DBP MAE (mmHg) | Location | N | Maneuver Required | Continuous Estimation |
| --- | --- | --- | --- | --- | --- | --- |
| This work [2024] | 5.31 ± 7.32 | 4.27 ± 2.35 | Chest/Arm | 16 | No | Yes |
| Carek et al. [2021] | 4.03 ± 4.75 | 2.24 ± 2.72 | Wrist | 21 | Yes | No |
| Liu et al. [2023] | 5.31 ± 4.89 | 3.57 ± 6.71 | Finger | 22 | No | No |
| He et al. [2022] | 6.18 ± 9.35 | 6.07 ± 6.13 | Wrist | 76 | Yes | No |

**Fig. 5.**
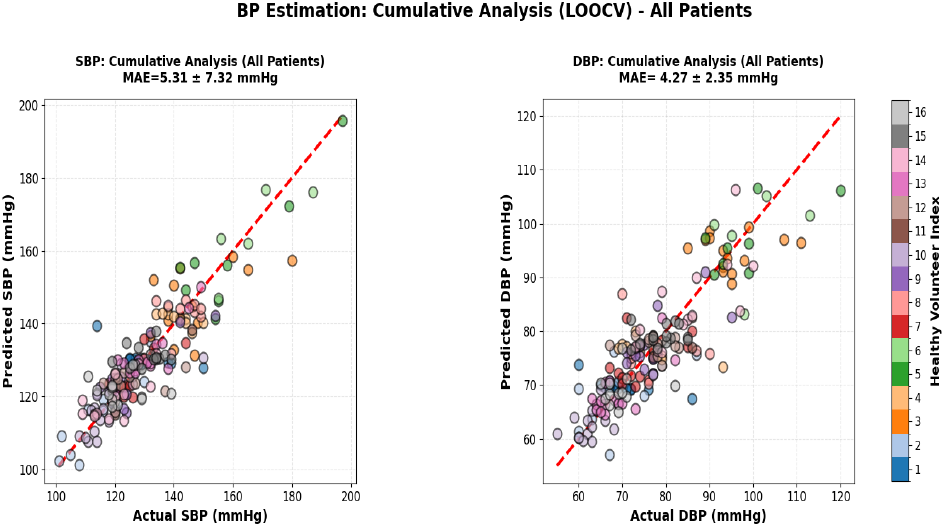
Cumulative leave-one-out cross-validation (LOOCV) analysis of blood pressure estimation across all patients.

**Fig. 6.**
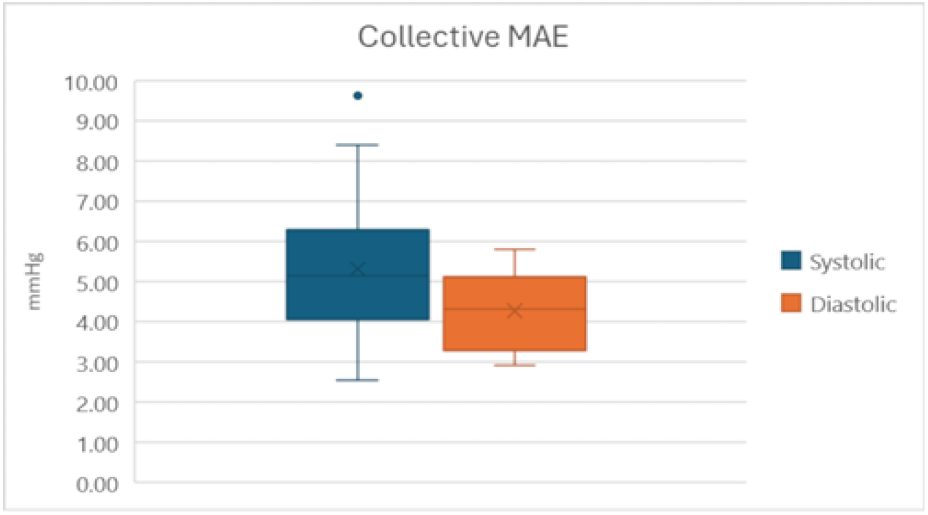
The distribution of predicted systolic and diastolic mean absolute error for all participants.

**Fig. 7.**
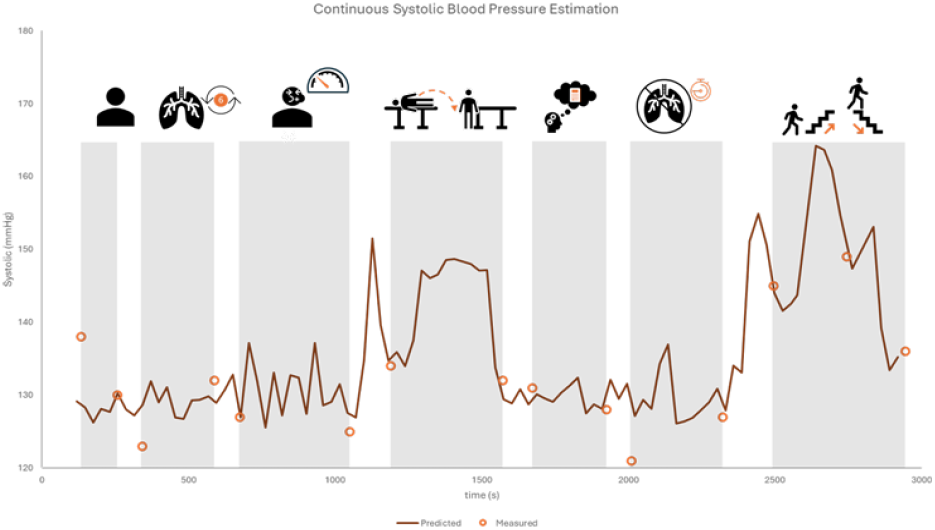
The results of continuous blood pressure estimation over the course of the 7 session research study.

Figure 8 illustrates the ensemble mean pre-ejection period (PEP) responses during the active standing session, grouped by blood pressure (BP) category. Non-hypertensive individuals exhibited a prominent and sustained increase in PEP following transition to standing, whereas hypertensive participants showed no such increase and demonstrated higher PEP variability throughout. The mean PEP difference before and after standing served as a robust discriminator between hypertensive and non-hypertensive groups, with linear separation for all but one participant; this exception, who had hypothyroidism, likely reflects altered myocardial contractility response. These findings highlight PEP’s value as a noninvasive physiological marker for distinguishing blood pressure status during an orthostatic transition.

**Fig. 8.**
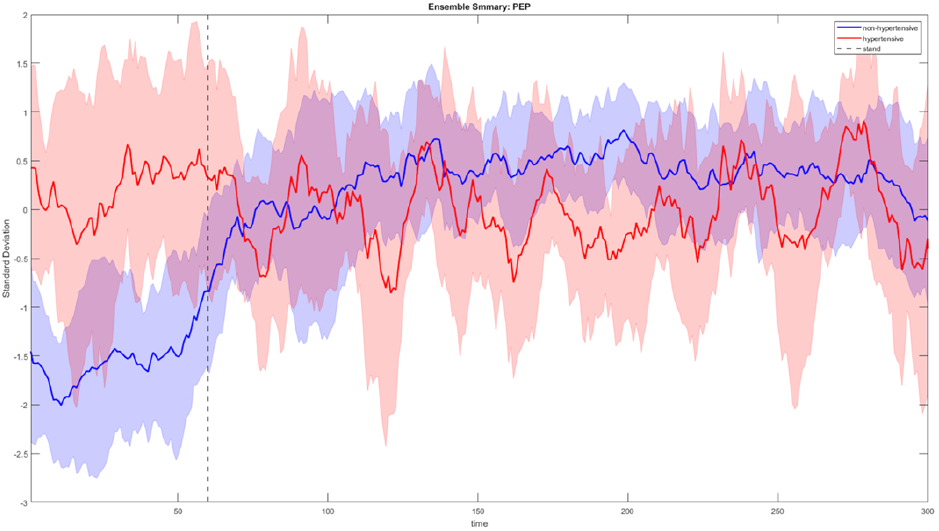
The mean PEP for all hypertensive (red) and non-hypertensive (blue) participants during the active stand test along with one standard deviation visualized by the shaded regions and the point of standing represented by the dashed line. Non hypertensive show an increase in PEP after standing while hypertensive participants exhibited a less observable change when transitioning from supine to standing.

## Conclusion

This work demonstrated the feasibility of blood pressure estimation during routine activities using a novel wearable system. The MOSAIC sensor confirmed that pulse arrival time-based measurements enable accurate, continuous blood pressure estimation during movement including orthostatic transitions and stair climbing without manual intervention. The MOSAIC system establishes a foundation for accurate non-invasive continuous blood pressure monitoring in active populations throughout their daily routines. As an alternative to conventional approaches such as blood pressure cuffs which interrupt daily routines and have low temporal resolution, the MOSAIC system offers a solution for providing the realtime data needed to reduce the widespread prevalence of mismanaged hypertension.

Device optimization remains essential for widespread adoption. Current limitations include battery size and power consumption, which restrict multi-day operation. Event-driven data collection could reduce energy use by capturing only physiologically meaningful periods. Device miniaturization may be achieved by replacing Ag/AgCl electrodes with flexible dry electrodes and interfacing directly with the NRF52832 microcontroller, enabling nearly a two-fold size reduction. Incorporating flexible materials and additional sensing modalities, such as sweat-based biochemical monitoring, offers further opportunities for enhancement. Realizing the system’s full potential requires real-time feedback to users or caregivers through either cloud-based processing or edge artificial intelligence. Cloud processing reduces on-device computation, increasing power efficiency and data collection bandwidth, but introduces latency unsuitable for critical healthcare applications. Conversely, embedded machine learning algorithms eliminate infrastructure dependence and latency, enabling fully standalone real-time health monitoring.

In conclusion, the research addresses the growing impact of hypertension through an innovative noninvasive wearable BP sensor capable of standalone real-health monitoring. By enabling continuous measurement outside of conventional clinical environments, this technology has the potential to significantly expand access to hypertension managements, particularly to populations with limited access to quality healthcare. Real-time availability allows faster and more informed treatment to adjustments, improving blood pressure control and reducing hypertension’s role as comorbid risk factor for stroke, heart-failure and other cardio-vascular disease. Overall, this work demonstrates the promise of distributed wearable sensor arrays to advance the next-generation health monitoring and to greatly improve outcomes for individuals living with hypertension.

## Data Availability

All data produced in the present study are available upon reasonable request to the authors

